# Cross-modality image translation of 3 Tesla Magnetic Resonance Imaging to 7 Tesla using Generative Adversarial Networks

**DOI:** 10.1101/2024.10.16.24315609

**Authors:** Eduardo Diniz, Tales Santini, Karim Helmet, Howard J. Aizenstein, Tamer S. Ibrahim

## Abstract

The rapid advancements in magnetic resonance imaging (MRI) technology have precipitated a new paradigm wherein cross-modality data translation across diverse imaging platforms, field strengths, and different sites is increasingly challenging. This issue is particularly accentuated when transitioning from 3 Tesla (3T) to 7 Tesla (7T) MRI systems. This study proposes a novel solution to these challenges using generative adversarial networks (GANs)—specifically, the CycleGAN architecture— to create synthetic 7T images from 3T data. Employing a dataset of 1112 and 490 unpaired 3T and 7T MR images, respectively, we trained a 2-dimensional (2D) CycleGAN model, evaluating its performance on a paired dataset of 22 participants scanned at 3T and 7T. Independent testing on 22 distinct participants affirmed the model’s proficiency in accurately predicting various tissue types, encompassing cerebral spinal fluid, gray matter, and white matter. Our approach provides a reliable and efficient methodology for synthesizing 7T images, achieving a median Dice of 6.82%,7,63%, and 4.85% for Cerebral Spinal Fluid (CSF), Gray Matter (GM), and White Matter (WM), respectively, in the testing dataset, thereby significantly aiding in harmonizing heterogeneous datasets. Furthermore, it delineates the potential of GANs in amplifying the contrast-to-noise ratio (CNR) from 3T, potentially enhancing the diagnostic capability of the images. While acknowledging the risk of model overfitting, our research underscores a promising progression towards harnessing the benefits of 7T MR systems in research investigations while preserving compatibility with existent 3T MR data. This work was previously presented at the ISMRM 2021 conference (Diniz, Helmet, Santini, Aizenstein, & Ibrahim, 2021).

## 1 INTRODUCTION

Magnetic Resonance Imaging (MRI) technology has offered high definition and noninvasive insights into the intricacies of human brain structure and function (Hagmann et al., 2006). This advancement has considerably improved our understanding of brain morphological and functional changes throughout life and in various neurodegenerative conditions (Ashburner & Friston, 2000). In addition, it has spearheaded the discovery of potential biomarkers for numerous neurological disorders (Ross et al., 2012), thereby opening the doors to early detection and therapeutic intervention.

The field of MRI has been shifting from conventional/clinical 3 Tesla (3T) MRI systems towards high-field strength 7 Tesla (7T) MRI systems (Uğurbil, 2014). The benefits of 7T MRI include augmented signal-to-noise ratio (SNR), enhanced visualization of brain structures due to reduced voxel size, and an improved contrast-to-noise ratio (CNR) resulting from more distinguishable tissue relaxation times (Uğurbil, 2014). Despite these advantages, increased costs, restricted availability, increased susceptibility artifacts, a higher specific absorption rate (SAR), and radiofrequency (RF) inhomogeneity may complicate image acquisition and interpretation (van Osch & Webb, 2014).

The field has seen significant advances in the design and application of RF coils that are instrumental in overcoming the challenges of 7T MRI, chiefly ensuring high-quality and high-resolution acquisition (Santini, Wood, et al., 2021). However, transitioning longitudinal patient studies from the widely used 3T systems to the more advanced 7T systems presents substantial hurdles. The primary reason for this is the innate differences in the images generated by the two systems, especially the contrast variations among the soft tissues. These may affect direct image comparisons and complicate longitudinal analyses (Obusez et al., 2018).

Other factors, such as variability in image quality and systematic differences between varying field strengths, can complicate statistical analyzes of aggregate data (Fortin et al., 2018). These difficulties underscore the need for efficient spatial adaptive data normalization methodologies capable of handling variances across different field strengths; they are paramount in strengthening our understanding of the neuroanatomical shifts and progressions associated with normal aging and pathological processes.

Artificial intelligence (AI) and machine learning technologies have significantly shaped numerous sectors, including healthcare (Bohr & Memarzadeh, 2020). In particular, medical imaging is being improved by the potential of deep learning methods, which extends their reach into the complex realm of neuroimaging (Shen, Wu, & Suk, 2017). Specifically, applying Convolutional Neural Networks (CNNs) in image classification tasks has improved diagnostic precision and efficiency (Litjens et al., 2017). Building on this momentum, Generative Adversarial Networks (GANs) and their variants have emerged as powerful tools for medical image analysis, including tasks such as image synthesis, segmentation, and data augmentation (Nie et al., 2018).

Introduced by Goodfellow et al., (2014), GANs comprise a unique model structure employing two deep neural networks: the generator and the discriminator. They engage in an adversarial process, aiming to generate synthetic data resembling the original distribution. These networks have demonstrated proficiency in modeling complex data distributions (Arjovsky, Chintala, & Bottou, 2017; Salimans et al., 2016), which is beneficial for various image transformation tasks.

CycleGANs, an extension of GANs, provide a solution to unpaired image-to-image translation tasks (Zhu, Park, Isola, & Efros, 2017), including photo enhancement (Chen & Koltun, 2017), style transfer (Gatys, Ecker, & Bethge, 2016), and image synthesis (T.-C. Wang et al., 2018). In particular, within the realm of medical imaging, its applications have been transformative. CycleGANs have also shown potential in tasks such as lesion synthesis (Guerrero et al., 2018). These networks can enrich rare disease datasets by generating synthetic lesion images, facilitating better disease detection and diagnosis. In addition, they have been used in tasks such as organ segmentation by transforming organ-specific images into simplified representations that facilitate the segmentation process (Cai, Zhang, Cui, Zheng, & Yang, 2019).

One significant application pertains to the translation of images in different modalities, such as converting Computed Tomography (CT) images to MRI images and vice versa (Wolterink, Leiner, Viergever, & Isgum, 2017). In practical scenarios, patients may not undergo CT and MRI scans due to cost, radiation exposure, or other considerations. By training in unpaired CT and MRI datasets, CycleGAN has demonstrated the capability to generate synthetic but anatomically accurate MR images from CT scans and, conversely, to produce CT-like images from MR scans.

Cross-modal image translation, defined by the machine learning and imaging community, consists of converting images from one modality to another, retaining the crucial content while modifying the style to resemble the target modality (Zhu et al., 2017). 3T and 7T, despite employing the identical principle of nuclear magnetic resonance, exhibit divergent operational mechanics due to distinct field strengths. This differentiation manifests itself in significant differences in image quality, resolution, CNR, susceptibility effects, and spectral separation (Vaughan et al., 2001). In this regard, 3T MRI can be considered one modality and 7T MRI another. The conversion process from 3T to 7T through GANs fulfills this criterion, as it retains the intrinsic information (content) from the 3T images and translates the image properties (style) to align with that of 7T MRI.

This study uses a dataset of unpaired 3T and 7T MR images to train a CycleGAN-based model capable of translating 3T MRI data into synthetic 7T data. This approach aims to address the problem of spatial adaptive MR data harmonization, allowing the combined analysis of cross-sectional and longitudinal data from 3T and 7T MRI systems.

## 2 MATERIALS & METHODS

### 2.1 Model Architecture

The landscape of machine learning has been significantly shaped by the emergence of GANs, as conceptualized by Goodfellow et al., (2014). This novel model structure, built on game-theoretic principles, employs two deep neural networks in strategic interaction: the generator and the discriminator. The generator aims to generate synthetic data replicating the actual data distribution, while the discriminator strives to discern the synthesized data from the real data. This adversarial dance evolves over iterative training sessions, yielding refined performance from both networks and improved proficiency in modeling complex data distributions (Arjovsky et al., 2017; Salimans et al., 2016).

Taking a step further, CycleGANs provide a robust solution to unpaired image-to-image translation, a challenging problem in computer vision. Zhu et al., (2017) introduced CycleGAN’s architecture, comprising two GANs containing a generator and a discriminator. The generator creates compelling images of the target domain, deceiving the discriminator, which aims to differentiate real images from the translated ones within the target domain. The design of CycleGAN is such that each generator maps an input image from a source domain to a target domain and vice versa to ensure cyclical consistency (Zhu et al., 2017).

A feature that distinguishes CycleGAN from traditional GANs is its cycle consistency loss. This loss function aims to ensure that the cycle of translating an image from one domain to another and back again reconstructs the original image. Let *X* denote the domain of images, and *Y* denote another domain of images. We can represent an image from *X* as *x ∈ X*, and an image from *Y* as *y ∈ Y*. Let us construct image datasets from each domain: *D*_*X*_ = {*x*_1_, *x*_2_, …, *x*_*m*_} consisting of *M* images from *X*, and *D_Y_* = {*y*_1_, *y*_2_, …, *y*_*n*_} consisting of *N* images from *Y*. Now, let the mappings *G* : **X* → Y* and *F* : *Y → X* be the generators between the two domains. Then, the cycle consistency loss is:

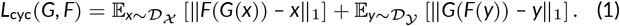

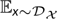 and 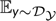 are the expectations taken over the datasets *D_X_* and *D_Y_*, respectively. The terms ∥*F* (*G*(*x*)) – *x*∥_1_ and ∥*G* (*F*(*y*)) – *y*∥_1_ measure the absolute difference between the original image and the image that has been translated to the other domain and back, *i*.*e*., *F* (*G*(*x*)) means *G* translates an image *x ∈ D*_*X*_ into the domain *Y* and then translated to *X* by *F*. Similarly, *G* (*F*(*y*)) takes an image *y ∈ D_Y_*, translates it into domain *X* by *F* and then converts it back to *Y* by *G*. The cycle consistency loss should be small if the model performs well, indicating that the original and the cycled-back image are nearly identical.

Our study uses a CycleGAN to deal with unpaired 3T and 7T MRI data. Figure 1 shows a schematic representation of the architecture and functioning of the CycleGAN model. Our implementation integrates two generator networks based on the U-Net architecture (Ronneberger, Fischer, & Brox, 2015), with residual blocks inserted between the encoding and decoding stages (He, Zhang, Ren, & Sun, 2016).

**FIGURE 1.**
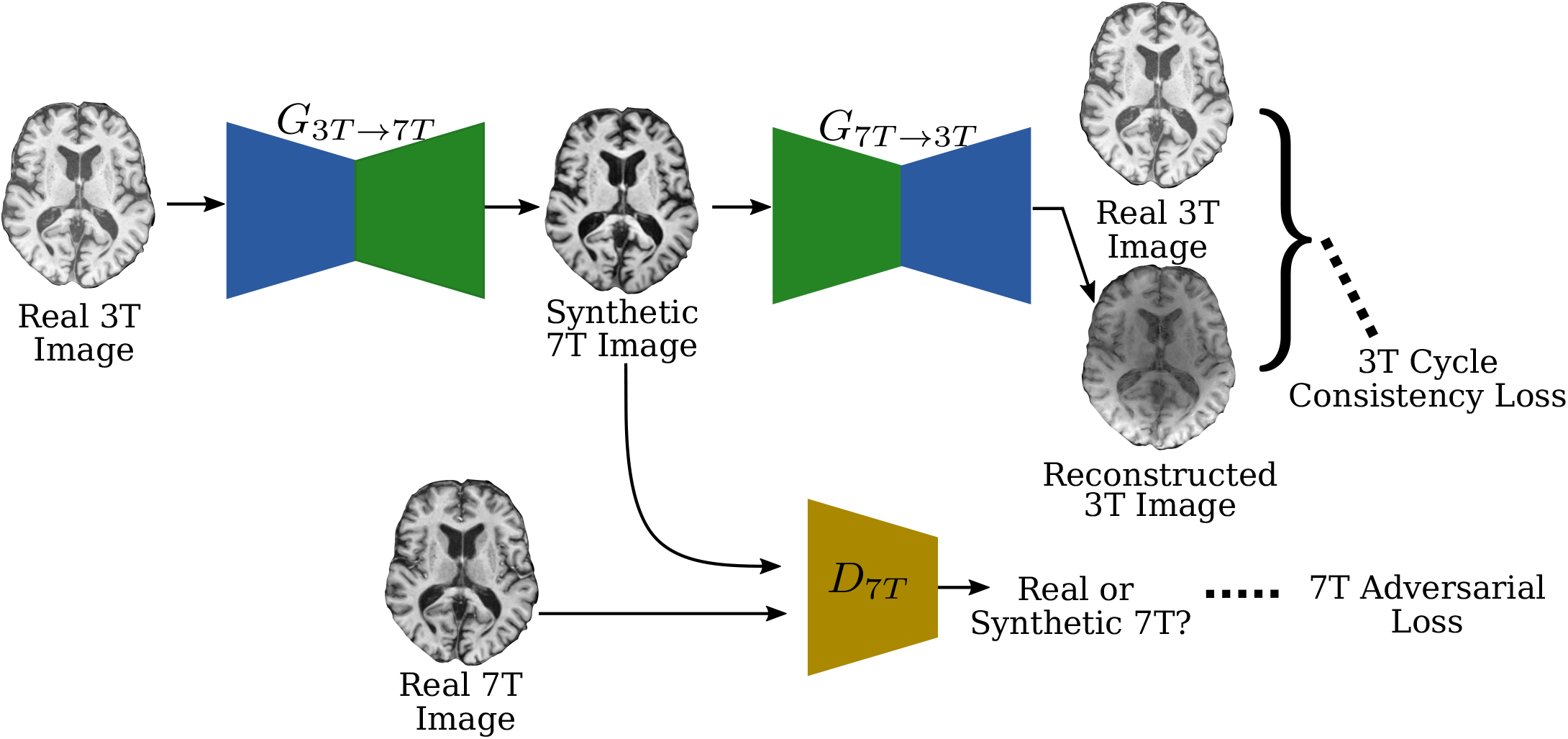
The cyclical consistency of the CycleGAN. This figure visually demonstrates the 3T cycle consistency. The 7T loop is conceptually symmetric. The generators are marked as performing the mappings *G*_3*T→*7*T*_ : 3*T →* 7*T* and *G*_7*T→*3*T*_ : 7*T →* 3*T*. The Real 3T image is fed to the *G*_3*T→*7*T*_ (top left), producing the synthetic 7T image (top middle). The *D*_7*T*_ discriminator differentiates between the Real and Synthetic 7T images and strives to minimize 7T adversarial loss (bottom left). The synthetic 7T image is fed into the *G*_7*T→*3*T*_ generator, which produces the reconstructed 3T image. The 3T cycle consistency loss ensures the 3T image can be reconstructed from the synthetic 7T image (top right).

U-Net is a CNN architecture renowned for its utility in biomedical image segmentation. It is named after its U-shaped structure, which consists of a contracting path to capture context and a symmetric expanding path for precise localization of features. The expanding pathway uses transposed convolutions, with skip connections that transfer feature maps from the contracting path to recover spatial information.

Residual blocks, proposed in the ResNet architecture by He et al., (2016) resolve training difficulties encountered with deep neural networks. These blocks consist of several convolutions, after which the input is added to the output, creating a residual connection. This configuration allows the network to learn residual functions concerning the layer’s inputs, facilitating the training of deeper models.

The model also uses instance normalization and stride-2 convolutions. Instance normalization is often used in style transfer tasks (Ulyanov, Vedaldi, & Lempitsky, 2016). It calculates the mean and variance for each instance separately, leading to the relative scaling of the activations. This technique is beneficial for disentangling content and style in image generation tasks. Stride-2 convolutions, frequently utilized in CNNs, help reduce the spatial dimensions of feature maps and allow the model to learn more abstract representations in deeper layers (Goodfellow, Bengio, & Courville, 2016).

Our CycleGAN model uses 70 *×* 70 PatchGANs for the discriminator networks. PatchGANs, as described by Isola et al., (2017), classify whether each patch in an image is real or fake instead of assessing the picture as a whole. This patch-level classification enables the generation of sharper and more contextually accurate images in tasks such as image-to-image translation.

### 2.2 Dataset

We leveraged an extensive collection of unpaired and paired 3T and 7T MR images from diverse sources to ensure a rich and comprehensive dataset. The 3T MRI data were primarily sourced from the Human Connectome Project (HCP), a globally recognized brain imaging dataset that contains high definition multimodal brain imaging data from healthy adults (Sotiropoulos et al., 2013). Specifically, structural 3T images were sourced from a subset of 1112 scans in the HCP’s 1200 subjects data release. The images were acquired using a Siemens 3T Connectome Skyra and a standard 32-channel Siemens receive head coil at Washington University in St. Louis. The scans were conducted using a T1-weighted (T1w) Magnetization Prepared RApid Gradient Echo (MPRAGE) protocol with a repetition time (TR) of 2400 *ms*, echo time (TE) of 2.14 *ms*, flip angle (FA) of 8^°^, field of view (FOV) of 224 *mm ×* 224 *mm*, and isotropic resolution of 0.7 *mm*, lasting approximately 8 minutes.

The inclusion criteria for the 3T HCP data required participants to be between 22 and 35 years old with no significant history of psychiatric disorder, substance abuse, neurological or cardiovascular disease and the ability to provide valid informed consent. Participants were required to have a Mini Mental Status Exam score above 28.

Exclusion criteria included multiple nonprovoked seizures or a diagnosis of epilepsy, genetic disorders such as cystic fibrosis, the use of prescription medications for migraines in the past 12 months, and conditions such as multiple sclerosis, cerebral palsy, brain tumor, stroke, sickle cell disease, thyroid hormone treatment in the past 12 months, current treatment for diabetes (excluding gestational or diet-controlled diabetes), head injury, and premature birth. Other exclusion criteria were the presence of unsafe metals or devices in the body (such as cardiac pacemakers, cochlear implants, aneurysm clips), current or historical use of chemotherapy or immunomodulatory agents that could affect the brain, pregnancy, and moderate to severe claustrophobia.

The 7T dataset was sourced from the 7TBRP dataset, acquired at the RF Research Facility at the University of Pittsburgh, a specialized research center for high-field imaging. The dataset included 490 subjects’ structural 7T MR images. The scans were acquired using a Siemens Magnetom 7T whole-body MR scanner and an in-house developed first generation of the Tic-Tac-Toe head radio-frequency coil system (Santini et al., 2018; Santini, Wood, et al., 2021; Krishnamurthy et al., 2019; Santini, Koo, et al., 2021). The scans were performed using a T1w MPRAGE protocol with a TR of 3000 *ms*, TE of 2.17 *ms*, bandwidth of 391 *Hz*/*Px*, GRAPPA reconstruction with acceleration factor *R* = 2, and isotropic resolution of 0.75 *mm*, lasting approximately 5 minutes.

The exclusion criteria for the 7TBRP dataset included pregnancy or lactation, acute medical problems that could result in neurocognitive or brain dysfunction, including diabetes mellitus, coronary artery disease, and causes cerebral vasculities, such as peripheral vascular disease. Other exclusion criteria were contraindications to MRI scanning, such as electronic implants, magnetically activated implants, tattoos above the shoulders, or brain implants. Data acquisition in all studies was performed following the protocol approved by local institutional review boards. All participants were older than 18 years and were able to provide their written informed consent prior to participation.

A set of paired (acquired in the same participants) 3T-7T data from the 7TBRP dataset was used to compare the synthetic 7T data from3T and the real 7T data. The data set consisted of MR images from 22 subjects. The study-specific inclusion and exclusion criteria for the paired data were similar to those for the unpaired 7T datasets.

### 2.3 Data Preprocessing

The first stage of pre-processing, bias field correction, was performed using FMRIB’s Automated Segmentation Tool (FAST) part of FSL (Y. Zhang, Brady, & Smith, 2001). This technique mitigates the intensity inhomogeneity observed within MR images due to receive coil profile and patient positioning differences (Vovk, Pernus, & Likar, 2007). If not correctly trained, deep learning models can learn from these inhomogeneities and become biased, leading to sub-optimal performance. Bias field correction can rectify this, enabling the model to concentrate on the salient features of the image (Arnold et al., 2001). This process results in enhanced feature extraction, improved segmentation, and registration results, which further enhances the effectiveness of subsequent processing and training phases (Tustison et al., 2010).

Following bias field correction, we performed spatial normalization using the Statistical Parametric Mapping (SPM) toolbox’s coregister function, co-registering the images via normalized mutual information to the Montreal Neurological Institute 152 standard (MNI152) space (Ashburner, 2007; Penny, Friston, Ashburner, Kiebel, & Nichols, 2011). Normalization facilitates the comparison and integration of data between subjects, as each voxel location corresponds to the same anatomical structure across all images (Evans, Janke, Collins, & Baillet, 2012). This alignment minimizes the need for the neural network to learn these invariances, potentially improving performance (Cao et al., 2017).

In the next stage, we transform the image intensities to a standard normal range of [–1, 1] using SPM’s image calculator function. This transformation minimizes intersubject variability, promoting improved image data consistency (Nyul & Udupa, 1999). Furthermore, intensity normalization allows the model to focus more on structural or semantic differences between images than on absolute pixel intensities (Goodfellow et al., 2016). This is critical during the early stages of training as it prevents the gradients from vanishing or exploding, resulting in a more stable optimization process and faster convergence (LeCun, Bottou, Orr, & Müller, 2002). Furthermore, intensity normalization can also make the learning process less dependent on the specific units used. This is especially important when combining data from multiple sources with different measurement units or scales (Bishop & Nasrabadi, 2006).

Finally, we skull-stripped the images using the Brain Extraction Tool (BET) within FSL, removing non-brain tissue from the MRI images and thereby reducing dimensionality, which can improve the efficiency of subsequent model training (Iglesias & Sabuncu, 2015; Smith, 2002). For computational efficiency, we transformed 3D images into 2D by extracting axial slices using FSL’s fslslice function, which is less computationally intensive while preserving critical anatomical information.

### 2.4 Model Training

Training a deep learning model requires orchestrating various strategies and methods to optimize performance. In the context of our CycleGAN model implemented within the TensorFlow framework (Abadi et al., 2016), we had to consider various elements such as computational hardware, optimization algorithms, weight initialization, regularization techniques, and learning rate schedules.

We used NVIDIA Titan RTX GPUs (24GB of memory) for training and adopted an iterative training strategy; the discriminators were updated four times for each update of the generators within the training loop. This prevented the generators from overpowering the discriminators, which is critical to supporting the dynamics of the adversarial process (Goodfellow et al., 2014).

We used early stopping as a regularization technique to control the model’s capacity and prevent overfitting. We stopped training when no significant improvement in the performance of the validation set was observed, *i*.*e*., the validation loss did not decrease by 0.1% over five consecutive epochs (Prechelt, 2002). Furthermore, L1/L2 regularization was employed to encourage the model to learn distributed and sparse representations, which can increase its generalizability (Ng, 2004).

We used the Adam optimizer (Kingma & Ba, 2014) for the optimization algorithm. Its adaptive learning rate and efficient computation make it a superior choice for training deep learning models, especially for achieving faster and more effective convergence. The Xavier weight initialization method was used for all convolutional layers (Glorot & Bengio, 2010). In addition, all bias terms were initialized to zero. Proper weight initialization can significantly improve the convergence rate and prevent problems such as vanishing or exploding gradients, making the optimization process more stable.

Finally, we incorporated learning rate annealing into the training process to fine-tune the speed at which the model learns. The learning rate was set at 10^−4^ for the first 100 epochs and was linearly decreased to zero. By gradually reducing the learning rate, the model can make significant updates to learn the gross structure of the data in the early stages and then fine-tune its weights in later stages, allowing for smoother and more effective convergence (Bengio, 2012).

### 2.5 Model Evaluation

The traditional approach to assessing GANs, primarily focusing on discriminator loss, needs an explicit measure of sample quality. This work thus evaluates the quality of training using a pre-trained deep neural network to embed the samples, inspired by Heusel et al., (2017) methodology. We adopted a ResNet-50 architecture pre-trained on ImageNet (He et al., 2016), given its effectiveness in similarity searching in the embedding space for various forms of imaging data, including MRI, even without fine-tuning (Yosinski, Clune, Bengio, & Lipson, 2014). In this context, the quality of the samples is evaluated on the basis of their realism and “closeness” to the target domain. We calculated the Fréchet ResNet Distance (FRD) and the Cosine ResNet Distance (CRD) between each sample in the ResNet-50 embedding space to facilitate a comparison between 3T and 7T MRI features.

Let *D*_*X′*_ = *G*(*D_Y_*) denote the dataset of synthetic samples and ResNet(*D_X_*) and ResNet(*D*_*X′*_) denote the ResNet-50 embeddings of real and synthetic samples, respectively. The FRD, calculated as the Fréchet Distance (Dowson & Landau, 1982), or, equivalently, the Wasserstein-2 distance (Vallender, 1974) between the ResNet(*D_X_*) and ResNet(*D*_*X ′*_) embeddings is given by:

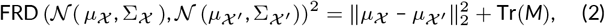

Where

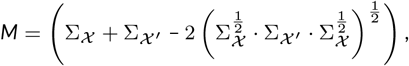

*N* (*µ_X_*, Σ_*X*_) and *N* (*µ*_*X ′*_, Σ_*X ′*_) are the multivariate Gaussian distributions over ResNet(*D_X_*) and ResNet(*D*_*X ′*_), respectively, ∥ *·* ∥_2_ represents the Euclidean norm, and Tr the trace.

Likewise, the CRD is the average cosine distance between each real

*z ∈* ResNet(*D_X_*) and synthetic *z*_*′*_ *∈* ResNet(*D*_*X ′*_):

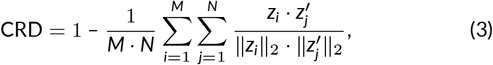

where *z*_*i*_ represents the *i*-th feature embedding in ResNet(*D_X_*) and *z*_*′*_ represents the *j*-th feature in ResNet(*D*_*X ′*_). *M* and *N* are the total real and synthetic samples, respectively. This expression calculates the cosine similarity between each pair of feature embeddings, normalizes it by their Euclidean norms, and then averages the distances over all pairs. Finally, we measured the sample quality over the synthetically generated 7T MR images. The accuracy of the generated synthetic 7T MR images was evaluated by cross-referencing these images with their respective real 7T ground truth counterparts. The images were segmented into white matter (WM), gray matter (GM), and cerebrospinal fluid (CSF) using FSL’s FAST segmentation tool. We then used two key and widely used evaluation metrics in medical imaging: the Dice Coefficient (Dice) and the Percentual Area Difference (PAD). These metrics were indispensable for evaluating the model’s performance and provided imperative insights for future model enhancement.

The Dice, introduced by Dice, (1945), measures the overlap between the predicted and the ground-truth segmentation. Mathematically, for an expected pixel set 𝒜 and the ground truth pixel set 𝔅, the Dice is:

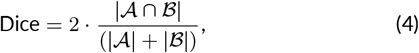

where |*·*| represents the cardinality of a set. The range of Dice ranges from 0 (indicating no overlap)to 1 (indicating a perfect overlap), making it an excellent indicator of the congruence between predicted and real segmentation (Zou et al., 2004).

Conversely, the PAD measures the discrepancy in the size between the predicted and the ground-truth segmentation. Specifically, it evaluates the model’s accuracy in estimating the size of the segmented objects. For an expected area *A* and the ground truth segmentation area *B*, the PAD is computed as:

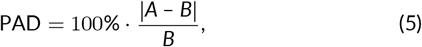

where |*·*| is the absolute value. The PAD is inversely proportional to the performance of the segmentation, with smaller values indicating a superior performance (Huang, Sun, Tseng, Li, & Qian, 2019).

## 3 RESULTS

The performance of the model was assessed using multiple parameters, including generators, discriminators, and cycle consistency losses in the validation dataset; FRD and CRD embedding quality measures; and a detailed analysis of the prediction for various brain tissue types and Dice and PAD sample quality metrics from independent test data.

### 3.1 Training Performance

We assessed the model’s performance by first analyzing the generator and discriminator losses in the validation dataset. The generator and the discriminator essentially participate in a minimax game, where the generator seeks to create images that the discriminator cannot distinguish from the real 7T images. In contrast, the discriminator aims to accurately classify real images from synthetic ones (Goodfellow et al., 2014).

Figures 2(a) and 2(b) show that the tug-of-war between generators and discriminators resulted in the generator losses gradually decreasing, demonstrating the generators’ increasing proficiency in creating images that the discriminators struggle to differentiate from real MRI images. In tandem, the discriminators’ losses also decreased, indicative of their improving ability to distinguish between real and generated images. At minima, the convergence of the losses substantiates the efficacy of the model in producing synthetic MRI-like images from real data.

**FIGURE 2.**
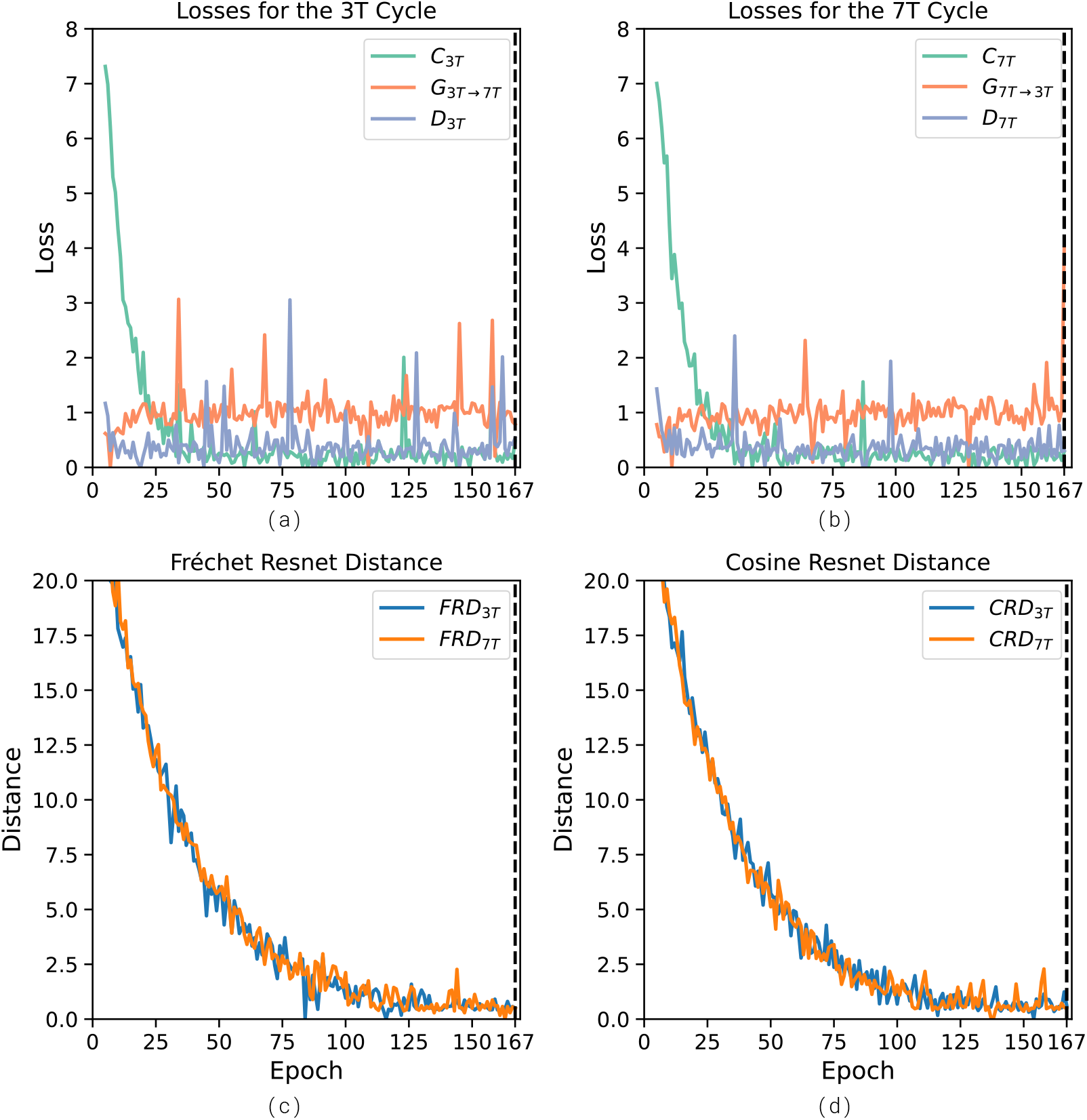
Progression of CycleGAN training dynamics and quality metrics for 3T to 7T MRI translation. (a) Generator, Discriminator and Cycle Consistency Losses for the 3T loop, (b) Generator, Discriminator, and Cycle Consistency Losses for the 7T loop, (c) Fréchet ResNet Distance (FRD), (d) Cosine Resnet Distance (CRD). The graphs illustrate the evolution of loss functions and quality metrics over training epochs. The vertical black dotted line mark epoch 167, where we halted training since we did not observe an improvement bigger than 0.1% in the 3T cycle consistency loss for five consecutive epochs.

However, this is evidence of improvement only during the early stages of training. Beyond that, since the generator and discriminator losses plateaued after the 14th epoch, just the cycle-consistency losses suggest that the output continued to improve. Hence, to quantitatively assess the progression of the CycleGAN, we apply the metrics FRD and CRD described in Methods 2.5 to the data generated at each training epoch. This strategy allows us to examine the training dynamics of the CycleGAN. As shown in Figures 2(c) and 2(d), FRD and CRD though slightly noisy, stop decreasing beyond the 125th epoch. In general, Figure 2 shows that the embedded MRI captures more relevant features in the data, with the measures of transformation quality showing a consistent and smooth decrease throughout the training process.

### 3.2 Visual Representation of Results

Further, an independent test dataset evaluated the generalization ability of the model, which involved using the model to translate 3T images from new subjects, unseen during training, into synthetic 7T images.

Figure 3 shows examples of real and synthetic 7T MRI image pairs generated by our model. The synthetic images manifest convincing resolution and contrast properties that mimic the attributes of real 7T images, demonstrating the model’s success in generating high-fidelity 7T-like images from 3T data.

**FIGURE 3.**
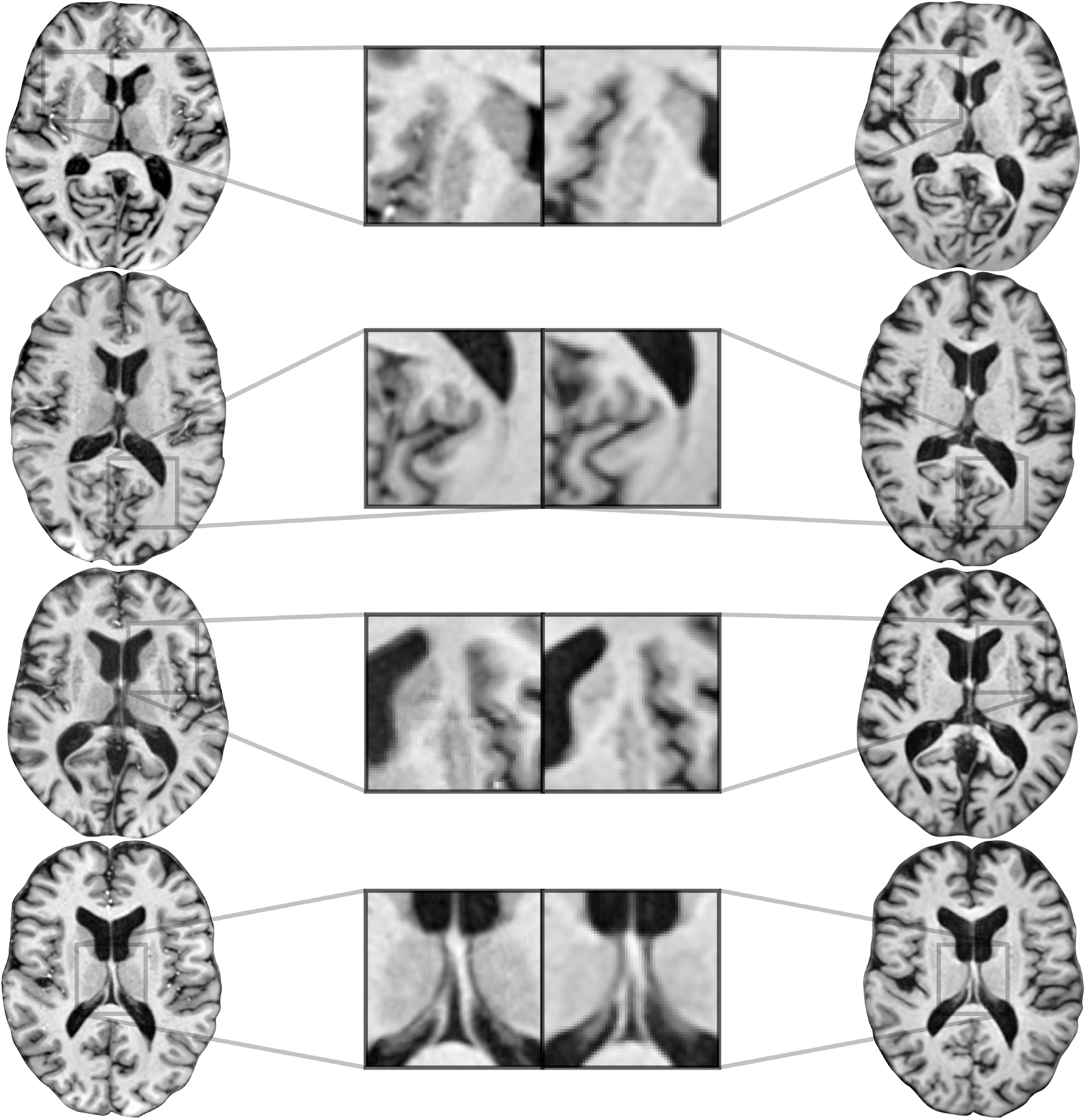
Examples of real 7T MR images (left column) and the corresponding synthetic 7T images (right column) generated by our model.

### 3.3 Tissue Type Specific Prediction

Diving deeper, we analyzed the performance of the model in generating specific brain tissue types—CSF, GM, and WM—by computing the Dice and the PAD. The results, visualized using box plots (Figure 4), revealed that the model achieved a median Dice of 83.62% for CSF, 81.42% for GM and 89.75% for WM. Concurrently, the median PAD was 6.82% for CSF, 7.63% for GM, and 4.85% for WM. These figures demonstrate consistent performance across all tissue types, corroborating the model’s robustness in translating detailed tissue-specific characteristics from 3T to 7T.

**FIGURE 4.**
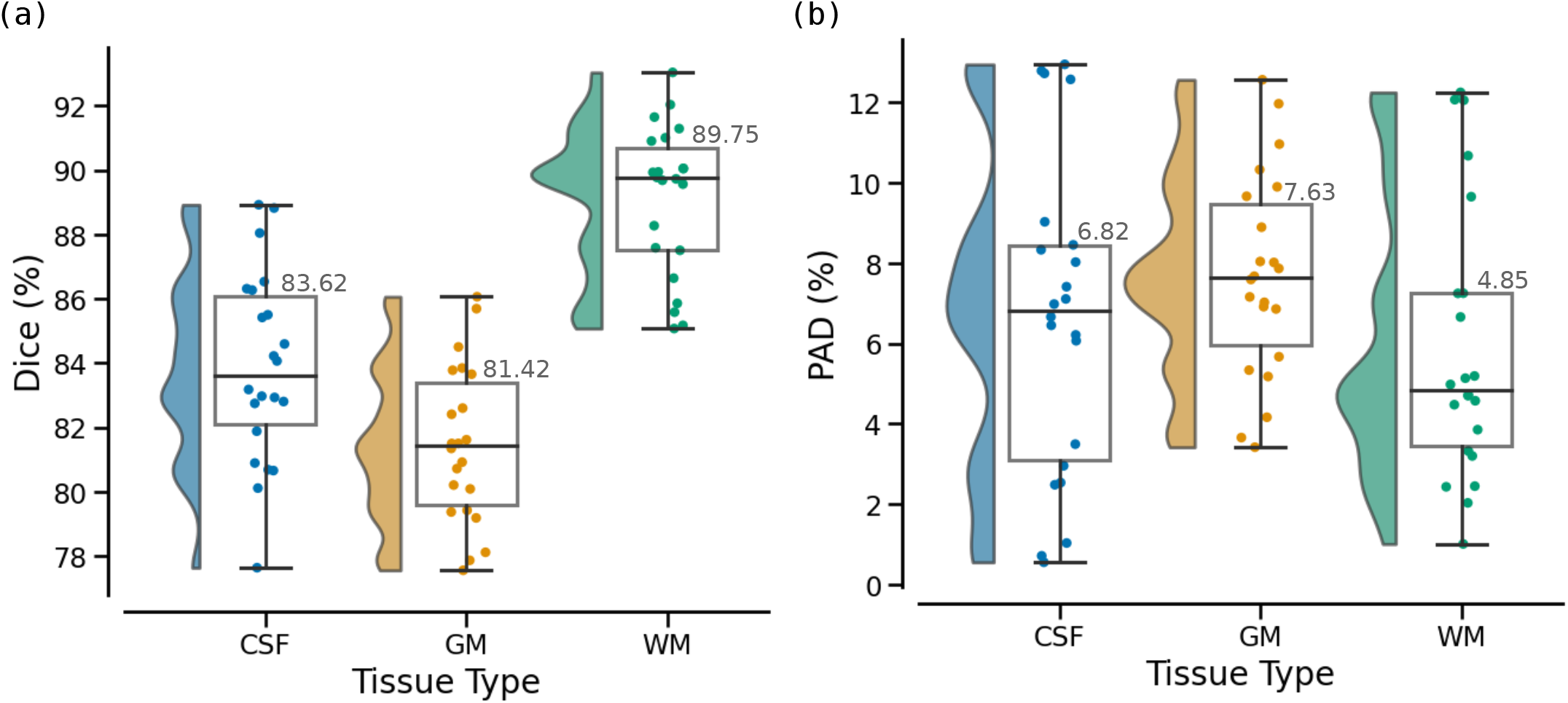
Group-level segmentation performance for the validation (22 subjects) data set across different tissue types—Cerebral Spinal Fluid (CSF), Gray Matter (GM), and White Matter (WM). We plot single patient data overlaid on group-level whisker box plots (center, median; box, 25th to 75th percentiles; whiskers, 1.5 interquartile range) and the smoothed data distribution. The median Dice for CSD, GM and WM are 83.62%, 81.42%, and 89.75%, respectively. The median percentual area difference (PAD) for CSF, GM, and WM are 6.82%, 7.63%, and 4.85%, respectively.

## 4 DISCUSSION

The present study sought to contribute with a novel approach to spatial adaptive MR data normalization between the 3T and 7T MR modalities, an increasing challenge in medical imaging. Using CycleGAN, an unsupervised generative adversarial network, our model demonstrated promising performance in generating clear 7T-like MR images from 3T inputs, evidenced by the high Dice and PAD(Taha & Hanbury, 2015; Zou et al., 2004). These scores affirm the model’s competence in maintaining the original morphological features of the brain and preserving the clinical relevance of the images, which has a significant implication in the advancement of neurological research (Moeskops et al., 2016; Ronneberger et al., 2015).

The successful performance of the model, especially in tasks that require global contrast properties, supports the hypothesis that our approach enables a seamless transition to 7T MR systems without jeopardizing the quality of previously obtained 3T data (Ashburner & Friston, 2000; Johansen-Berg & Behrens, 2006). This holds promise for facilitating consistent and robust data analysis, mitigating potential bias and loss of statistical power associated with data from difference MRI field strengths in longitudinal studies (Schaer et al., 2008; Little & Rubin, 2019).

Previous research efforts have probed into creating 7T images using advanced deep-learning methodologies (Hou et al., 2016; Qin et al., 2019). However, our model stands out due to its unsupervised learning aspect and its ability to function without resorting to information in the frequency domain (Klosowski & Frahm, 2017; Huang et al., 2019). The effectiveness of the CycleGAN model in minimizing image variance underlines its potential to tackle the challenge of cross-modality image translation between different MRI field strengths (Adriany et al., 2008; Van der Velden et al., 2015).

Our study offers a promising approach to cross-modality MR image translation, allowing the efficient utilization of 3T and 7T MRI technologies in longitudinal studies examining brain health and pathology (Uğurbil, 2014; Keuken & Forstmann, 2015). The strategy we propose constitutes a progress towards exploiting the capabilities of cutting-edge neuroimaging technologies while preserving the value of existing imaging data. This spatial adaptive normalization tactic helps bridge the technological gap, potentially accelerating the identification and validation of imaging biomarkers for neurological conditions (Bourgeat et al., 2015; W. Zhang et al., 2015).

Our research has made a significant step forward in advancing the application of CycleGAN to transform 3T to 7T MR data. However, the success of the translation task is highly dependent on various factors, including the quality and nature of the input data, the model architecture, and the training procedure. Despite our progress, it is essential to consider the following factors as potential limitations that can impact the results.

One of the primary limitations concerns the 2D nature of the implemented model. Adopting a 2D CycleGAN model was primarily due to computational considerations; however, the inherent lack of a threedimensional context by independently processing each slice results in slice-to-slice inconsistencies (Çiçek, Abdulkadir, Lienkamp, Brox, & Ronneberger, 2016; Milletari, Navab, & Ahmadi, 2016). Furthermore, patch-based image processing can result in loss of local spatial information, which is critical in medical image segmentation (Tajbakhsh et al., 2020). These inconsistencies may impact the quality and fidelity of the synthesized 7T images. As Yushkevich et al., (2006) have demonstrated, 3D analysis can significantly enhance image interpretation and extract critical information from complex structures. Although implementing 3D CycleGAN architecture would alleviate this concern, the extensive computational demand often hinders its deployment.

The second potential pitfall is associated with an intrinsic property of CycleGANs, namely the non-bijective or many-to-many mapping between domains (Zhu et al., 2017). This can lead to struggles with controlling the mode of output (Ghosh, Kulharia, Namboodiri, Torr, & Dokania, 2018) and non-functional transformations, which introduce potentially unrealistic information, or “hallucinations,” into the synthetic images (Almahairi, Rajeshwar, Sordoni, Bachman, & Courville, 2018). These limitations and the difficulty of maintaining the semantic consistency of anatomical structures in synthesized images (Chartsias, Joyce, Giuffrida, & Tsaftaris, 2018) hinder their diagnostic applicability (G. Wang et al., 2020). Consequently, future research should focus on devising strategies to rectify this challenge, such as regularizing the model with a perceptual loss (Johnson, Alahi, & Fei-Fei, 2016) or improving training procedures to facilitate a better generalization (Roth et al., 2020).

Variations in image quality across different MRI scanners and imaging protocols can also influence our model’s performance (Jovicich et al., 2006; Kruggel, Turner, Muftuler, & Alzheimer’s Disease Neuroimaging, 2010). This issue is frequently encountered in multi-center studies and can generate images that do not accurately represent real-world variance, reducing its practical use. Therefore, future research is warranted to assess the model’s resilience against such variations and verify its performance on diverse datasets, including those featuring pathological changes (van Opbroek, Ikram, Vernooij, & de Bruijne, 2015).

The question of data dependence and the potential for error in translation must also be mentioned. The translation’s success relies heavily on the quality of the input data, meaning that errors during preprocessing, like inaccurate skull-stripping or normalization, could adversely impact the output (Iglesias & Sabuncu, 2015). Similarly, the translation process could inadvertently introduce or exaggerate noise or artifacts not present in authentic 7T images, leading to misleading interpretations (Chen & Koltun, 2017).

The performance evaluation of segmentation algorithms plays a significant role in medical imaging. It determines the efficacy and reliability of these algorithms in clinical practice. We employed the Dice and PAD scores. Despite its widespread usage, the Dice score has a few limitations. One major drawback is that it needs to account for the geometric correspondence between the predicted and ground truth segments (Taha & Hanbury, 2015). It does not consider how well the predicted segment’s shape, location, and orientation match the ground truth. In some scenarios, this information is critical, mainly when the size and position of the segment are clinically significant (*e*.*g*., tumor segmentation) (Crum, Camara, & Hill, 2006).

Conversely, the PAD evaluates the size estimation accuracy of segmentation results. While PAD effectively measures size discrepancies, it does not provide information on the spatial overlap between the predicted and the ground truth segmentation. Thus, two segmentations with the same area but in different locations would yield a zero PAD, failing to account for the spatial mismatch (Isensee, Kickingereder, Wick, Bendszus, & Maier-Hein, 2017). This limitation is critical in situations where the accurate localization of the object of interest is essential.

### 4.1 Conclusion and future work

Our study underscores the transformative potential of blending advanced machine learning techniques with clinical applications, particularly within neuroimaging (Obermeyer & Emanuel, 2016; Esteva et al., 2019). Our results provide a powerful testimony to the potential of CycleGAN in handling unpaired data, thus offering a compelling solution to MR translation across different field strengths.

However, despite its merits, the CycleGAN model has limitations, and these challenges delineate promising avenues for future enhancement. Our study used generators based on the U-Net architecture (Ronneberger et al., 2015), which have delivered excellent results in medical image segmentation tasks. However, U-Net-based models primarily exploit local dependencies in the data, potentially neglecting long-range relationships between pixels. In contrast, with the advent of Vision Transformers (ViTs), there is potential to enhance our model’s performance further (Dosovitskiy et al., 2020). ViTs can capture these intricate, long-range dependencies, which could be highly advantageous for tasks involving complex morphological structures such as brain MR images (T.-C. Wang et al., 2018). This, in turn, could improve the segmentation performance, particularly for brain images that exhibit non-local relationships between anatomical structures (Petit et al., 2021).

## ACKNOWLEDGMENTS

The National Institutes of Health supported this work under award numbers: R01MH111265, R01AG063525, R01MH076079, and RF1AG025516. This research was partly supported by the University of Pittsburgh Center for Research Computing, RRID:SCR_022735, through the resources provided. Specifically, this work used the H2P cluster, supported by NSF award number OAC-2117681. The Human Connectome Project, WU-Minn Consortium partly provided 3T data (Principal Investigators: David Van Essen and Kamil Uğurbil; 1U54MH091657) funded by NIH and Washington University. CNPq partially supported Eduardo Diniz under the award: Ph.D. Scholarship Ciência sem Fronteiras, Brazil (#210150/2014-9).

## DATA AVAILABILITY

The trained model and code are available upon request.

## SUPPORTING INFORMATION

Additional supporting information may be found in the online version of the article at the publisher’s website.

